# Comparative analysis of symptom profile and risk of death associated with infection by SARS-CoV-2 and its variants in Hong Kong

**DOI:** 10.1101/2023.05.25.23289996

**Authors:** Kin On Kwok, Wan In Wei, Edward B. Mcneil, Arthur Tang, Julian Wei Tze Tang, Samuel Yeung Shan Wong, Eng Kiong Yeoh

**Author notes:** Joint first author (Kin On Kwok, Wan In Wei). Joint senior author (Samuel Yeung Shan Wong, Eng Kiong Yeoh).

## Abstract

**Introduction:** The recurrent multi-wave nature of COVID-19 necessitates updating its symptomatology. Before the omicron era, Hong Kong was relatively unscathed and had a low vaccine uptake rate among the old-old, giving us an opportunity to study the intrinsic severity of SARS-CoV-2 variants. A comparison of symptom patterns across variants and vaccination status in Hong Kong has yet to be undertaken. The intrinsic severity of variants and symptoms predictive of severe outcomes are also understudied as COVID-19 evolves. We therefore aim to characterize the effect of variants on symptom presentation, identify the symptoms predictive and protective of death, and quantify the effect of vaccination on symptom development.

**Methods:** With the COVID-19 case series in Hong Kong from inception to 25 August 2022, an iterative multi-tier text-matching algorithm was developed to identify symptoms from free text. Cases were fully vaccinated if they completed two doses. Multivariate regression was used to measure associations between variants, symptom development, death and vaccination status. A least absolute shrinkage and selection operator technique was used to identify a parsimonious set of symptoms jointly associated with death.

**Results:** Overall, 70.9% (54450/76762) of cases were symptomatic. We identified a wide spectrum of symptoms (n=102), with cough, fever, runny nose and sore throat being the most common (8.16-47.0%). Intrinsically, the wild-type and delta variant caused similar symptoms, with runny nose, sore throat, itchy throat and headache more frequent in the delta cohort; whereas symptoms were heterogeneous between the wild-type and omicron variant, with seven symptoms (fatigue, fever, chest pain, runny nose, sputum production, nausea/vomiting and sore throat) more frequent in the omicron cohort. With full vaccination, omicron was still more likely than delta to cause fever. Fever, blocked nose and shortness of breath were robustly jointly predictive of death as the virus evolved. Number of vaccine doses required for reduction in occurrence varied by symptoms.

**Discussion:** This is the first large-scale study to evaluate the changing symptomatology by COVID-19 variants and vaccination status using free-text reporting by patients. We substantiate existing findings that omicron has a different clinical presentation compared to previous variants. Syndromic surveillance can be bettered with reduced reliance on symptom-based case identification, increased weighing on symptoms robustly predictive of mortality in outcome prediction, strengthened infection control in care homes through universal individual-based risk assessment to enable early risk stratification, adjusting the stockpile of medicine to tally with the changing symptom profiles across vaccine doses, and incorporating free-text symptom reporting by patients.

## INTRODUCTION

### Evolving SARS-CoV-2 epidemiology in Hong Kong

Hong Kong has experienced five waves of Coronavirus Disease 2019 (COVID-19), caused by severe acute respiratory syndrome coronavirus 2 (SARS-CoV-2) [1]. The first two waves were driven by the ancestral wild-type virus, the third and the fourth were dominated by lineages B.1.1.63 and B.1.36.27, respectively, and the early fifth wave was mainly caused by the omicron subvariant BA.2 [1]. Hong Kong was relatively unscathed in the first four waves, which can be attributed to a hospital-based approach under which all confirmed or suspected cases were isolated. Before the fifth wave, Hong Kong was characterized by a low proportion (18%) of the old-old completing a two-dose primary vaccination series and predominantly vaccine-induced immunity at the population level [2]. To cope with the surge in cases during the fifth wave, an online declaration system was established for the community to self-report their infection and symptoms [3]. Against this background, the disease progression in Hong Kong enables revisiting the clinical spectrum of SARS-CoV-2 of primary infection episodes across vaccination groups amid a changing landscape of viral pathogenicity [4].

### Importance of identifying the symptom profile of SARS-CoV-2

The recurrent multi-wave nature of COVID-19 [5] makes it worth updating the symptomatology to improve disease recognition, particularly when it involves the telltale signs of severe health outcomes. As of 26 March 2023, weekly hospitalizations by COVID-19 exceeded that of influenza by 13-fold in the United Kingdom [6]. Though symptoms triggered by respiratory illnesses are generally alike [7], the persistent clinical sequelae after the acute phase of infection [8] and the complexity of symptoms added by the omicron variant [9] highlight the need to differentiate COVID-19 from other respiratory diseases. While the circulating omicron variant may induce less severe symptoms than its preceding counterparts [10], this benignity can be counterbalanced by an increased incidence of infection disrupting society via sickness. In fact, 46% of the patients infected with omicron regarded their illnesses as moderate or severe [11].

### Research gaps

Despite the importance of updating symptom profiles and disease burden as SARS-CoV-2 evolves, there are **four** research gaps that need addressing.

**First**, symptom profiles of COVID-19 vary across regions [12], but to our knowledge, we are not aware of any study comparing symptom patterns of infection across variants and vaccination status in Hong Kong. Such a clinical spectrum was studied initially [13,14], but the varying pathogenicity of SARS-CoV-2 [4] highlights the need to revisit the clinical presentation. Even under the same viral condition, symptom profiles vary with the host properties and the immunity level of the affected population, which is an interplay between natural infection and vaccine regimes (for example, the type of vaccine administered and timing of vaccination).

**Second**, the intrinsic severity of SARS-CoV-2 variants are understudied and most published studies have returned mixed findings. Intrinsic-severity estimates are indispensable information to plan upcoming infection control strategies against the public burden caused by immune-evasive pathogens, compromising the effectiveness of vaccination. On one hand, the intrinsic severity is less for omicron according to Nyberg and colleagues who reported a larger reduction (comparing omicron with delta) in the risk of hospitalization and death in unvaccinated cases than all cases [15], and by Zhang and colleagues who reported no significant difference in the blood profiles of the fully vaccinated and the non/partially vaccinated [16]. On the other hand, Whitaker and colleagues found that omicron cases were more likely to report cold-like symptoms [17]. Therefore, it is important to unravel the intrinsic severity of the concurrent and recent predominant SARS-CoV-2 variant, especially when the mainstream vaccine administered in Hong Kong has low efficacy against symptomatic infection of past strains [18], and when it takes time for the bivalent vaccine to be taken by the majority of the community.

**Third**, symptoms predictive of severe health outcomes are understudied as COVID-19 evolves. To our knowledge, such information was studied in the wild-type era by Vahey and colleagues who reported that vomiting, dyspnea, altered mental status, dehydration, and wheezing were significantly associated with hospitalization [19]. However, differences in replication and pathogenicity among SARS-CoV-2 variants [4] may present varied symptom-based predictors of mortality. By identifying which symptoms are predictive of severe outcomes, patients can be better triaged for medical procedures and prioritization of testing.

**Fourth**, established work based on structured responses of a predefined scope of symptoms might underestimate the prevalence and diversity of symptoms. For example, Vihta and colleagues recorded 12-16 symptoms [20] versus 32 preset symptoms in the ZOE app [21]). In addition, Malden and colleagues showed that free-text data of electronic medical records increased the identification of more cases for each symptom by 29-64% [22].

### Study aims and significance

In light of the above research gaps, this study makes use of free-text symptom data self-reported by cases to **(i)** characterize the (intrinsic and post-vaccination, wherever applicable) effect of variants on symptom presentation, **(ii)** identify the symptoms which are predictive and protective of death across variants, and **(iii)** quantify the effect of vaccination on symptom development. Following the World Health Organization’s end of the “public health emergency of international concern” declaration for COVID-19 [23], future infection control will focus on elderly and high-risk groups such that further investigations into the telltale signs of COVID-19 (as it evolves) will guide public health policies during patient triage.

## METHODS

### Data curation

We analyzed the case series of COVID-19 infections provided by the Centre for Health Protection (CHP) between 23 January 2020 and 25 August 2022 from which the following information was available: age, sex, chronic diseases, case classification (local or imported), date of case reporting to the CHP, symptoms experienced at the date of reporting, date of symptom onset, and vaccination date and type. The symptom data was initially solicited by nurse interviews, which was replaced with self-reporting on 26 February 2022.

### Iterative process of symptom extraction from free-text data

Symptoms were recorded in the form of free text in addition to four hard-coded symptoms (fever, chest pain, shortness of breath and breathing difficulty). Through compiling frequent lay-person descriptions by cases, a list was created to map their responses to standardized variables created based on the national ambulatory medical care survey (NAMCS) [24]. Classification by NAMCS was based on patients’ expression of their symptoms serving as the input to health services, which fits the current context of COVID-19 cases reporting their symptoms. To increase the accuracy of symptom identification, an iterative multi-tier text-matching algorithm was developed **(Table S1)**. A sample of 200 randomly selected cases was manually reviewed for symptom extraction to assess the accuracy of this algorithm.

### Classification of variants

Cases reported on or before 30 June 2020 were regarded as wild-type cases [1]. Starting from the third wave (1 July 2020), cases not sequenced for the determination of the variant or failed/had insufficient viral load for sequencing were unclassified. For the remaining cases, the variant was determined based on the results of whole genome sequencing by CHP upon positive results of polymerase chain reaction or rapid antigen testing.

### Definition of vaccination status

Cases were defined as unvaccinated if they did not receive any dose of vaccine on or before the symptom onset date (for symptomatic individuals) or the report date (for asymptomatic individuals). Cases were considered to complete a vaccine dose if that dose was received at least 14 days before the symptom onset date (for symptomatic individuals) or the report date (for asymptomatic individuals). Cases were defined as fully vaccinated if they completed two vaccine doses. A 14-day window period was assumed by convention as the human immune system takes time to build up protection [25].

### Statistical analysis

Cases with incomplete data were excluded from the analysis (**Figure 1**). Descriptive statistics, including proportions with binomial confidence intervals, were used to summarize the data. Multivariate regression models were fit to examine **(i)** the effect of variants on symptom development, **(ii)** the association of symptoms on elderly deaths, and **(iii)** the effect of vaccination on symptom development. The intrinsic severity of COVID-19 variants among elderly cases, measured as the occurrence of death within 28 days of symptom onset, was compared with Cox’s proportional hazards regression. All models were adjusted for age, sex, and reporting delay, defined as the time elapsed between symptom onset and the report date for symptomatic cases. When considering the overall effect regardless of variants, the analyses were also adjusted for variants. Adjusted odds ratios (aOR) and hazard ratios with 95% confidence intervals (CI) were presented visually.

**Figure 1.**
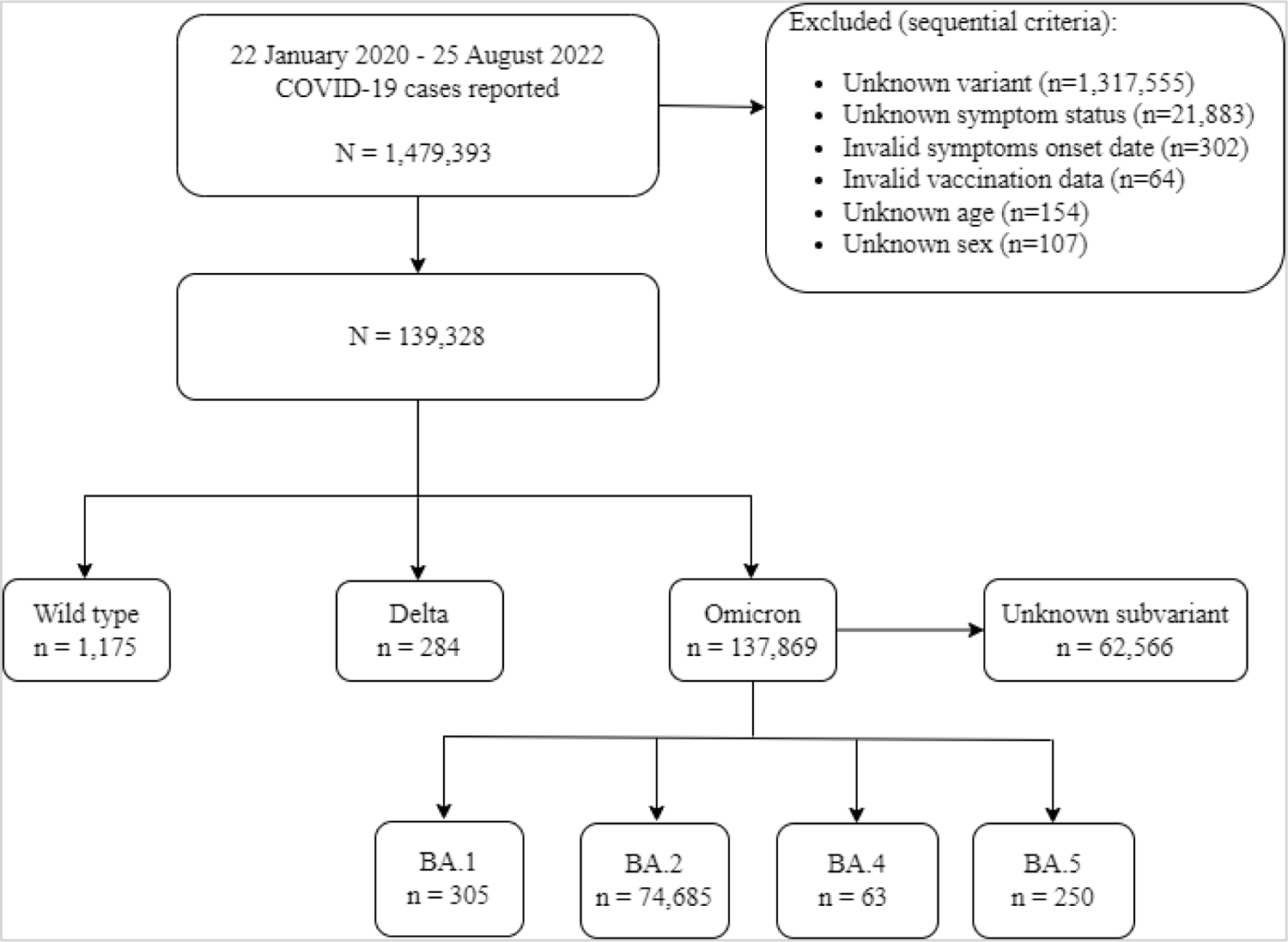
Study flowchart

We used a least absolute shrinkage and selection operator (LASSO) technique under the generalized linear regression framework to identify parsimonious symptoms jointly associated with death. The *glmnet* package in R was used to fit these models [26]. The value of the regularization parameter was determined using *k*-fold cross validation. The performance of these regression models was evaluated using the area under the receiver operating characteristic curve (AUC). All analyses were performed in R (version 4.2).

## RESULTS

### Subject characteristics

As of 25 August 2022, there were 1,479,393 COVID-19 cases reported in Hong Kong, among which 76,762 were included in the analysis with the majority being BA.2 cases (**Figure 1**). The reporting period of these cases ranged from 23 January 2020 to 19 July 2022. The study population consisted largely of adults aged 20-59 years (74.2%), followed by the elderly aged ≥60 years (15.3%) and young adults or minors (10.5%) (**Table 1**). The variant-specific proportion of local cases differed, ranging from over 95% for omicron (BA.2) and delta to 34.5% for wild type. All 1,175 wild-type cases were recorded on or before 30 June 2020, while the variants were mostly recorded in 2022. All wild-type cases were unvaccinated, but the circulation of its variants or sub-variants was amid an increasing level of vaccine-induced population immunity.

**Table 1.** Demographic and epidemiological characteristics of all included COVID-19 cases.

| Characteristic | Overall<br>N = 76,762 | Wild type<br>N = 1,175 | Delta<br>N = 284 | Omicron<br>(BA.1)<br>N = 305 | Omicron (BA.2)<br>N = 74,685 | Omicron<br>(BA.4)<br>N = 63 | Omicron<br>(BA.5)<br>N = 250 |
| --- | --- | --- | --- | --- | --- | --- | --- |
| Age group (years) |  |  |  |  |  |  |  |
| 0-9 | 4,397 (5.7) | 33 (2.8) | 12 (4.2) | 9 (3.0) | 4,332 (5.8) | 0 (0.0) | 11 (4.4) |
| 10-19 | 3,649 (4.8) | 172 (14.6) | 17 (6.0) | 19 (6.2) | 3,415 (4.6) | 4 (6.3) | 22 (8.8) |
| 20-29 | 11,785 (15.4) | 280 (23.8) | 35 (12.3) | 77 (25.2) | 11,337 (15.2) | 14 (22.2) | 42 (16.8) |
| 30-39 | 17,046 (22.2) | 249 (21.2) | 29 (10.2) | 63 (20.7) | 16,645 (22.3) | 3 (4.8) | 57 (22.8) |
| 40-49 | 15,015 (19.6) | 134 (11.4) | 44 (15.5) | 71 (23.3) | 14,704 (19.7) | 13 (20.6) | 49 (19.6) |
| 50-59 | 13,148 (17.1) | 146 (12.4) | 33 (11.6) | 38 (12.5) | 12,876 (17.2) | 13 (20.6) | 42 (16.8) |
| 60-69 | 7,463 (9.7) | 109 (9.3) | 46 (16.2) | 20 (6.6) | 7,263 (9.7) | 10 (15.9) | 15 (6.0) |
| 70+ | 4,259 (5.5) | 52 (4.4) | 68 (23.9) | 8 (2.6) | 4,113 (5.5) | 6 (9.5) | 12 (4.8) |
| Sex |  |  |  |  |  |  |  |
| Female | 40,968 (53.4) | 543 (46.2) | 154 (54.2) | 155 (50.8) | 39,954 (53.5) | 27 (42.9) | 135 (54.0) |
| Male | 35,794 (46.6) | 632 (53.8) | 130 (45.8) | 150 (49.2) | 34,731 (46.5) | 36 (57.1) | 115 (46.0) |
| Wave |  |  |  |  |  |  |  |
| 1-2 | 1,175 (1.5) | 1,175 (100.0) | 0 (0.0) | 0 (0.0) | 0 (0.0) | 0 (0.0) | 0 (0.0) |
| 5 | 75,587 (98.5) | 0 (0.0) | 284 (100.0) | 305 (100.0) | 74,685 (100.0) | 63 (100.0) | 250 (100.0) |
| Classification <sup>a</sup> |  |  |  |  |  |  |  |
| Imported <sup>b</sup> | 3,265 (4.3) | 770 (65.5) | 14 (4.9) | 304 (99.7) | 2,061 (2.8) | 30 (47.6) | 86 (34.4) |
| Local | 73,497 (95.7) | 405 (34.5) | 270 (95.1) | 1 (0.3) | 72,624 (97.2) | 33 (52.4) | 164 (65.6) |
| Vaccination status <sup>c</sup> |  |  |  |  |  |  |  |
| Unvaccinated | 14,001 (18.2) | 1,175 (100.0) | 95 (33.5) | 38 (12.5) | 12,677 (17.0) | 3 (4.8) | 13 (5.2) |
| One complete dose | 4,453 (5.8) | 0 (0.0) | 31 (10.9) | 8 (2.6) | 4,411 (5.9) | 0 (0.0) | 3 (1.2) |
| One incomplete dose | 2,002 (2.6) | 0 (0.0) | 6 (2.1) | 0 (0.0) | 1,996 (2.7) | 0 (0.0) | 0 (0.0) |
| Two complete doses | 38,425 (50.1) | 0 (0.0) | 130 (45.8) | 183 (60.0) | 38,057 (51.0) | 11 (17.5) | 44 (17.6) |
| One complete and one incomplete dose | 1,473 (1.9) | 0 (0.0) | 1 (0.4) | 0 (0.0) | 1,472 (2.0) | 0 (0.0) | 0 (0.0) |
| Three complete doses | 12,898 (16.8) | 0 (0.0) | 15 (5.3) | 69 (22.6) | 12,578 (16.8) | 48 (76.2) | 188 (75.2) |
| Two complete and one incomplete doses | 3,510 (4.6) | 0 (0.0) | 6 (2.1) | 7 (2.3) | 3,494 (4.7) | 1 (1.6) | 2 (0.8) |
| Presence of symptoms |  |  |  |  |  |  |  |
| No | 22,312 (29.1) | 302 (25.7) | 72 (25.4) | 131 (43.0) | 21,690 (29.0) | 32 (50.8) | 85 (34.0) |
| Yes | 54,450 (70.9) | 873 (74.3) | 212 (74.6) | 174 (57.0) | 52,995 (71.0) | 31 (49.2) | 165 (66.0) |

<sup>a</sup> Local cases consist of "local cases", "locally acquired cases", "possibly local cases", and "close contact of (possibly) local cases". Others were grouped as imported cases.
<sup>b</sup> The majority of imported cases were from the United Kingdom (13.4%), the United States (7.7%), and Singapore (4.4%).
<sup>c</sup> A complete dose refers to receipt of a vaccine dose given at least 14 days before the symptom onset date (for symptomatic cases) or the report date (for asymptomatic cases).

### Prevalence of symptoms

The majority (70.9%) of the cases were symptomatic (**Table 1**). Compared with the hard-coded symptoms, extraction from free-text data identified an additional 5336/11343 (47.0%) cases with fever, 714/2173 (32.9%) cases with shortness of breath or breathing difficulty, and 68/1770 (3.8%) with chest pain. Overall, a wide spectrum of 102 symptoms was identified, among which 30 symptoms had an overall prevalence of at least 0.5% **(Table S3)**. The free-text extraction showed that cases used varied expressions to describe their symptoms in the throat (for example, dry throat, itchy throat, sore throat, throat discomfort, throat irritation, tonsillitis) and in the muscle pain category (for example, bodyache, back pain, low extremity pain, unspecified muscle pain) **(Table S2)**. Regardless of variants and vaccination status, the most common symptoms were cough (range: 22.5-47.0%), fever (17.9-37.2%), runny nose (8.16-33.1%), sore throat (15.0-29.1%) and headache (4.96-14.6%) (**Figure 2**). Among the wild-type cases, three more symptoms stood out: disturbance of taste and/or smell (6.38%), shortness of breath (5.19%) and diarrhea (4.94%).

**Figure 2.**
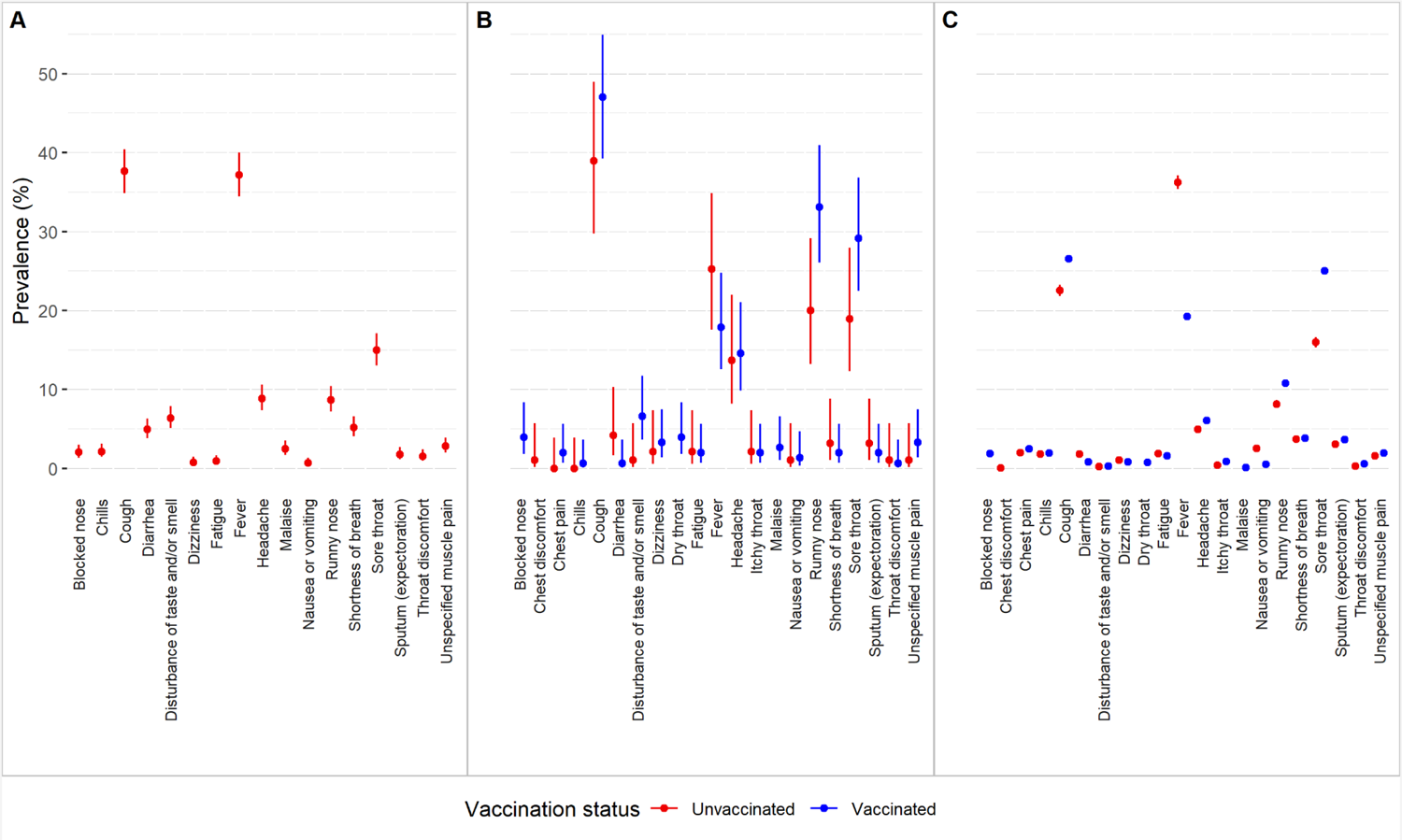
Prevalence of commonly reported symptoms among COVID-19 cases stratified by vaccination status for (A) wild-type, (B) delta and (C) omicron BA.2 variants. Only symptoms with a variant-specific combined (both the vaccinated and unvaccinated) prevalence >1% are shown.

### Effect of variant on symptom presentation

Effects of variant on symptom presentation were examined among the unvaccinated symptomatic cases (**Figure 3**), implicating the intrinsic severity of the variants in context. Omicron was more likely than wild-type to be associated with fatigue (aOR: 2.83 [95% CI: 1.53-5.21]), fever (aOR: 1.96 [1.69-2.27], chest pain (aOR:7.21 [2.96, 17.6]), runny nose (aOR: 1.36 [1.09-1.69]), expectoration (aOR: 2.54 [1.62-3.99], nausea/vomiting (aOR:5.13 [2.53-10.39]) and sore throat (aOR: 1.68 [1.41-2.00], but less likely to be associated with malaise, blocked nose, cough, diarrhea, throat discomfort, taste/smell disturbances, and headache. On the other hand, the symptoms associated with delta and the wild-type were more alike in that delta was more likely to be associated with only four symptoms: runny nose (aOR: 4.15 [2.30-7.49]), sore throat (aOR: 1.94 [1.08-3.48]), itchy throat (aOR: 36.58 [3.20-418.78]) and headache (aOR: 2.41 [1.24-4.68]). Differences in symptom presentation were also noted between omicron and delta for fever, runny nose, and headache.

**Figure 3.**
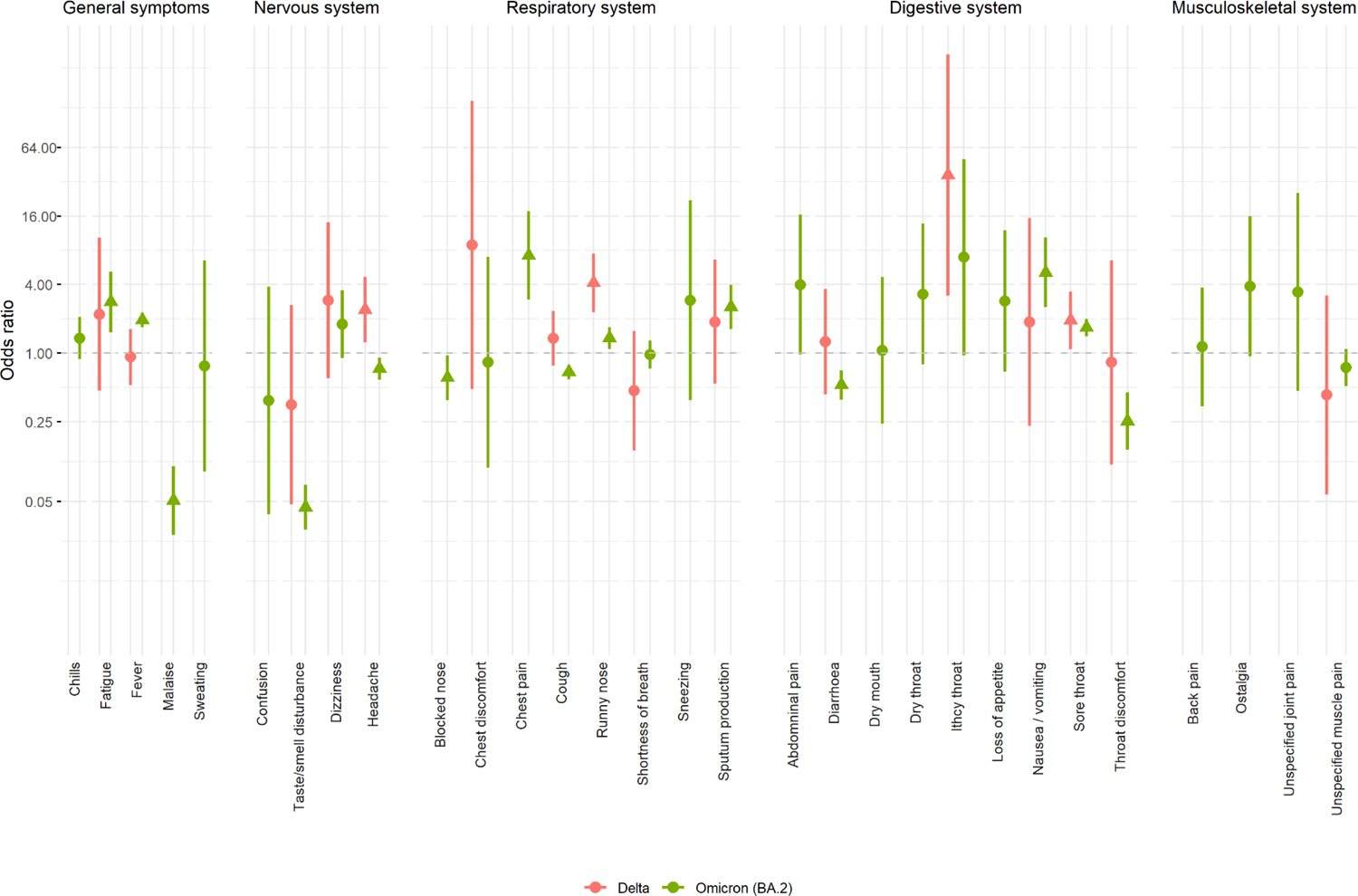
Effect of variant on symptom development among unvaccinated symptomatic cases (adjusting for age, sex, and reporting delay). Estimates that are significant and not significant are represented by solid triangles and circles, respectively.

Effects of variant on symptom presentation were examined among the fully vaccinated symptomatic cases (**Figure 4**). Omicron was more likely than delta to be associated with fever (aOR:1.66 [1.08-2.57]), but less likely to be associated with malaise, cough, runny nose, dry throat, taste/smell disturbances, dizziness, chest discomfort, palpitations (not shown in Figure 4) and headache.

**Figure 4.**
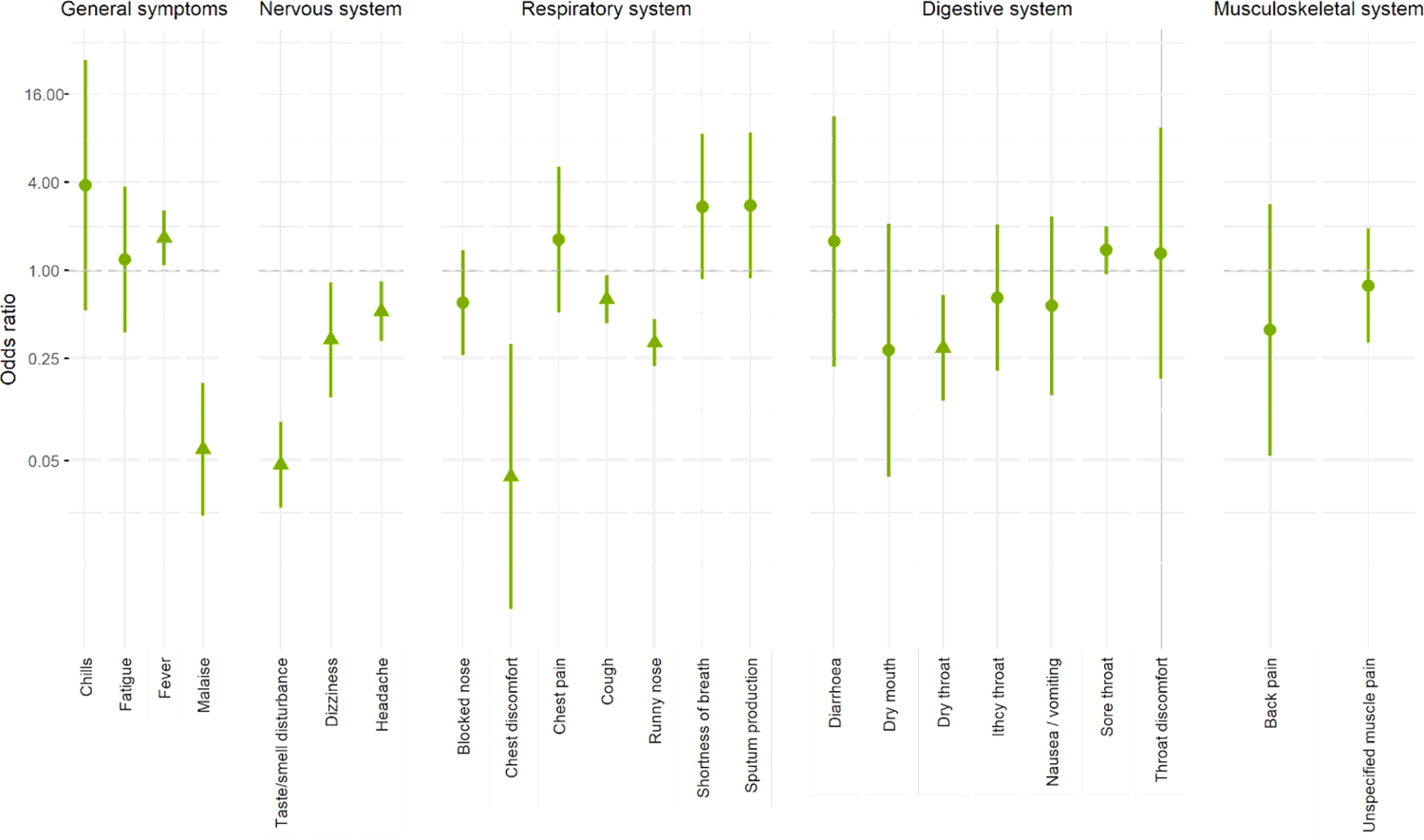
Effect of the omicron variant compared to delta on symptom development among fully vaccinated symptomatic cases. Solid triangles and circles represent estimates that are significant and not significant. (adjusting for age, sex, and reporting delay).

### Association between symptom presentation and death

The overall case fatality rate (CFR) was 0.25% (195/76,762), and that among the unvaccinated and the fully vaccinated were 1.11% and 0.05% respectively. Intrinsically, the hazard of death (within 28 days of symptoms onset) due to delta and omicron infections among elderly were similar, and were both higher than that attributable to the ancestral wild-type **(Figure S1)**. Among 75 decedents who were unvaccinated and symptomatic, 18 symptoms were ever reported: fever (64.0%) being the most common, followed by cough (37.3%), shortness of breath (25.3%) and sore throat (10.7%) (**Table 2**). Fever (aOR: 2.04 [1.22, 3.47]) and shortness of breath (aOR:2.25 [1.21, 4.03]) were predictive of death; whereas cough (aOR:0.51 [0.30, 0.84]) and sore throat (aOR:0.43 [0.19, 0.87]) were protective.

**Table 2.** Association of symptoms with death among unvaccinated symptomatic cases

| Symptom | Frequency |  |  | Logistic regression |  |  |
| --- | --- | --- | --- | --- | --- | --- |
|  | Overall<br>(N=7,988) | Alive<br>(N=7,913) | Dead<br>(N=75) | OR | 95% CI |  |
| General symptoms |  |  |  |  |  |  |
| Chills | 258 (3.2%) | 257 (3.2%) | 1 (1.3%) | 0.68 | 0.04 | 3.31 |
| Fatigue | 251 (3.1%) | 249 (3.1%) | 2 (2.7%) | 0.58 | 0.09 | 2.03 |
| Fever | 5,080 (63.6%) | 5,032 (63.6%) | 48 (64.0%) | 2.04 | 1.22 | 3.47 |
| Malaise | 41 (0.5%) | 40 (0.5%) | 1 (1.3%) | 2.06 | 0.11 | 12.0 |
| Nervous system |  |  |  |  |  |  |
| Disturbance of taste and/or smell | 104 (1.3%) | 103 (1.3%) | 1 (1.3%) | 5.85 | 0.31 | 31.6 |
| Dizziness | 145 (1.8%) | 144 (1.8%) | 1 (1.3%) | 0.78 | 0.04 | 4.07 |
| Headache | 758 (9.5%) | 756 (9.6%) | 2 (2.7%) | 0.39 | 0.06 | 1.33 |
| Respiratory system |  |  |  |  |  |  |
| Blocked nose | 139 (1.7%) | 138 (1.7%) | 1 (1.3%) | 3.67 | 0.20 | 19.1 |
| Chest pain | 262 (3.3%) | 261 (3.3%) | 1 (1.3%) | 0.43 | 0.02 | 2.13 |
| Cough | 3,350 (41.9%) | 3,322 (42.0%) | 28 (37.3%) | 0.51 | 0.30 | 0.84 |
| Runny nose | 1,162 (14.5%) | 1,157 (14.6%) | 5 (6.7%) | 0.53 | 0.18 | 1.25 |
| Shortness of breath | 538 (6.7%) | 519 (6.6%) | 19 (25.3%) | 2.25 | 1.21 | 4.03 |
| Sputum (expectoration) | 417 (5.2%) | 415 (5.2%) | 2 (2.7%) | 0.38 | 0.06 | 1.28 |
| Digestive system |  |  |  |  |  |  |
| Abdominal pain | 66 (0.8%) | 65 (0.8%) | 1 (1.3%) | 4.28 | 0.21 | 27.6 |
| Nausea or vomiting | 332 (4.2%) | 327 (4.1%) | 5 (6.7%) | 2.04 | 0.64 | 5.31 |
| Sore throat | 2,232 (27.9%) | 2,224 (28.1%) | 8 (10.7%) | 0.43 | 0.19 | 0.87 |
| Musculoskeletal system |  |  |  |  |  |  |
| Back pain | 30 (0.4%) | 29 (0.4%) | 1 (1.3%) | 14.2 | 0.73 | 86.0 |
| Unspecified muscle pain | 235 (2.9%) | 234 (3.0%) | 1 (1.3%) | 0.71 | 0.04 | 3.42 |

Remarks: All models were adjusted for age, sex, subvariant, and reporting delay.

Older age (≥60 years) significantly increased the odds of death for all symptoms (not shown in Table 2). With most decedents being elderly, we searched for a robust set of symptoms which were jointly predictive of death (**Table 3**). Overall, abdominal pain, blocked nose, malaise, shortness of breath, and fever were predictive of death while protective symptoms included cough, diarrhea, chest pain, voice disorders, sore throat, and loss of appetite. Among omicron cases, the predictive set included only blocked nose, shortness of breath, and fever. The predictive power of these sets of symptoms was good (AUC: 0.926).

**Table 3.**
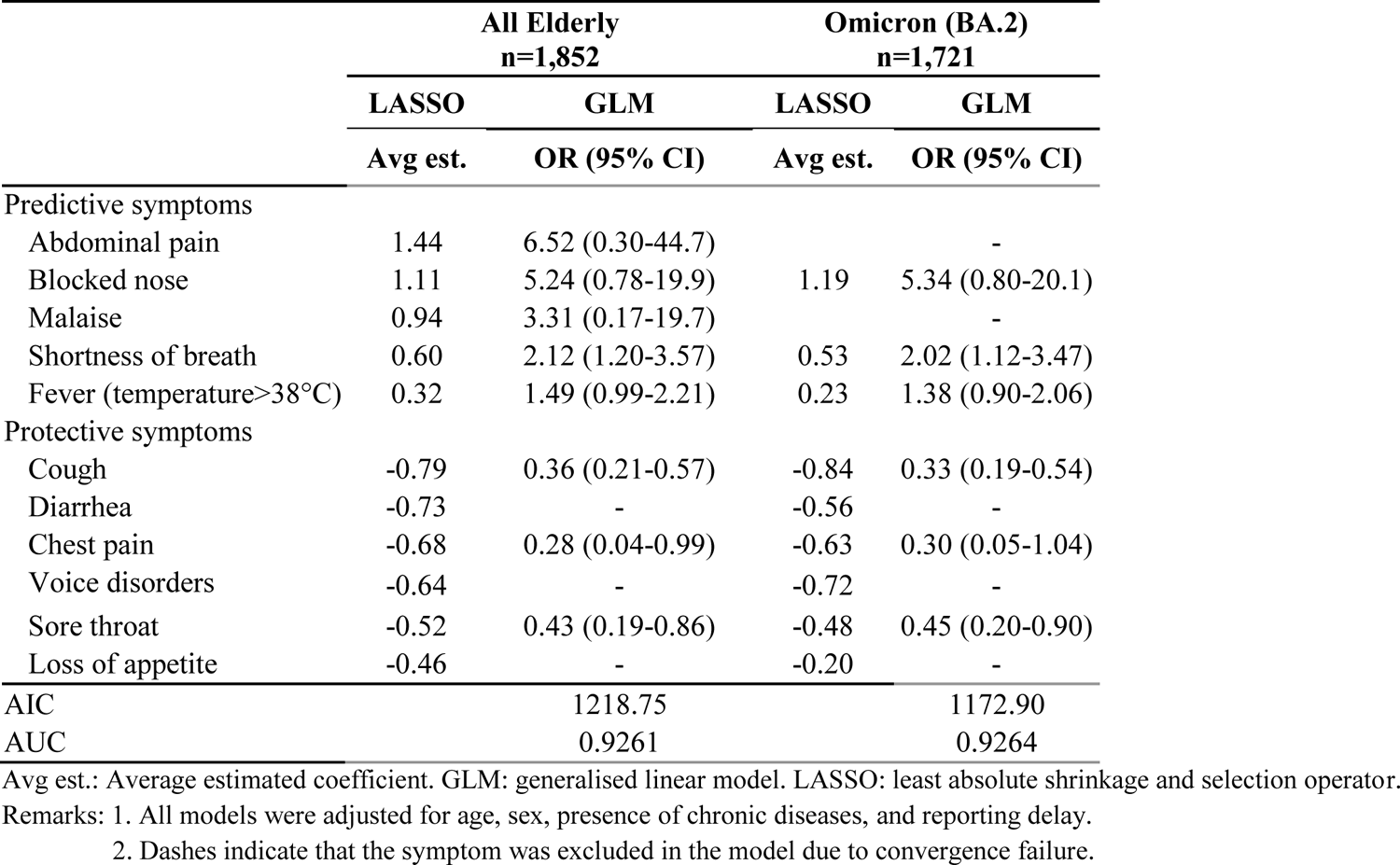
Average penalized LASSO estimates for symptoms associated with death and stepwise generalized linear models among unvaccinated elderly cases

### Effect of vaccination on symptom presentation

Considering the difference in symptom presentation between the young (<60 years) and the elderly (≥60 years) **(Figure S2)**, and older age is a significant risk factor for death, the extent of vaccination alleviating symptom presentation was examined among symptomatic elderly cases (**Figure 5**). Receipt of vaccination did not necessarily reduce the occurrence of symptoms, and in fact was associated with an increased chance of developing sore throat and some respiratory symptoms such as blocked nose, cough, and runny nose. The number of doses required to reduce individual symptoms also varied, from one dose for reducing fever, to two doses for reducing shortness of breath, to three doses for reducing chills. We also explored the effect of time elapsed since last vaccination on the occurrence of symptoms predictive of death (**Figure 6**). Compared with the number of doses, time elapsed since vaccination given the same number of doses did not alter the chance of occurrence of the predictive symptoms.

**Figure 5.**
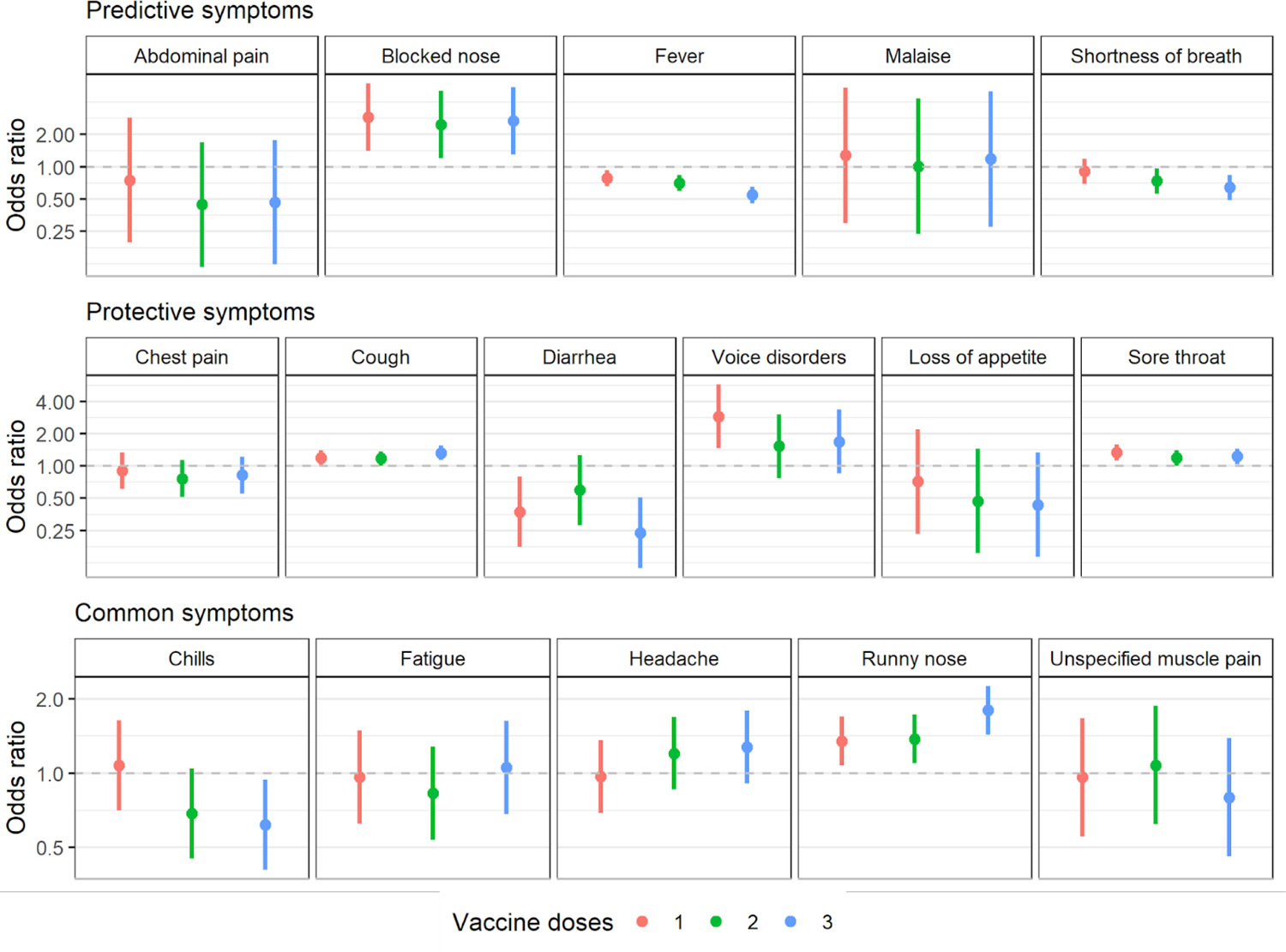
Effect of the number of vaccine doses on symptom development among symptomatic elderly cases, adjusting for age, sex, variant, and reporting delay. The reference group is the unvaccinated cases.

**Figure 6.**
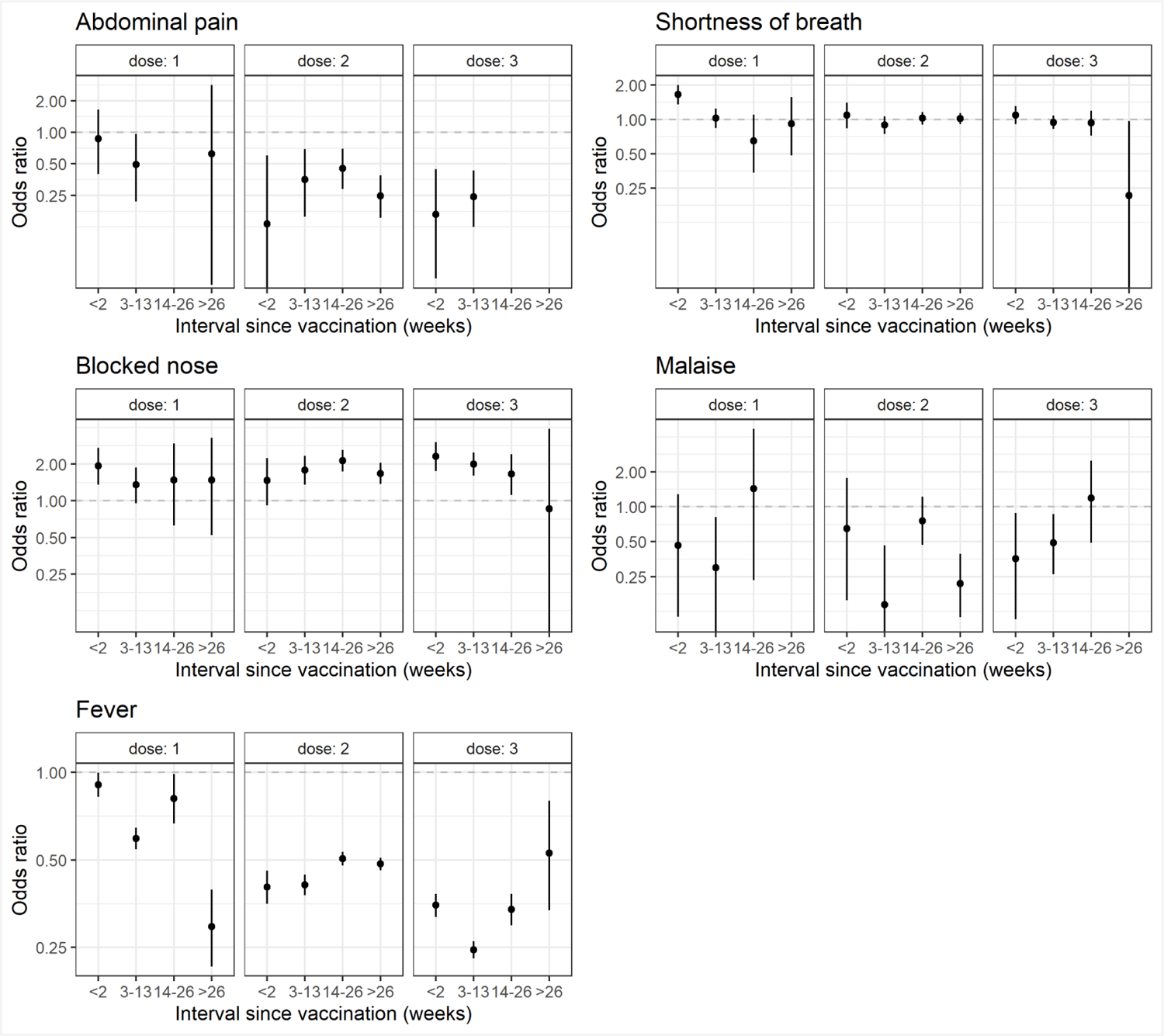
Effect of the time elapsed since the last vaccination on symptom development among symptomatic elderly cases, adjusting for age, sex, variant, and reporting delay. The reference group is the unvaccinated cases.

## DISCUSSION

### Principal findings

This is the first large-scale study to evaluate variations in symptom presentation by COVID-19 variants and vaccination status using free-text reporting by patients. We substantiate existing findings that omicron has a different clinical presentation compared to previous variants [27]. We identified a wide spectrum of symptoms (n=102) associated with SARS-CoV-2 infection, with the most common ones being cough, fever, runny nose and sore throat (8.16-47.0%). Intrinsically, symptoms caused by the wild-type and delta were generally similar, except that runny nose, sore throat, itchy throat and headache were more frequent in the delta cohort; whereas symptoms were heterogeneous between the wild-type and omicron, with seven symptoms (fatigue, fever, chest pain, runny nose, sputum production, nausea/vomiting and sore throat) more likely to be experienced by the omicron cohort, and seven symptoms (malaise, blocked nose, cough, diarrhea, throat discomfort, taste/smell disturbances, and headache) less likely to occur. Under full vaccination coverage, omicron was still more likely than delta to cause fever. Furthermore, fever, blocked nose and shortness of breath were robustly predictive of death even when the wild-type virus evolved to the omicron variant. Though the death rate was much lower in the fully vaccinated cohort, vaccination does not necessarily reduce the occurrence of symptoms, and the number of doses required to reduce symptoms varied by individual symptoms.

### Result implications

Our findings have five public health implications.

**First,** the weighing of symptom-based predictions of COVID-19 should be reduced. We found a high prevalence of upper respiratory tract infections among the predominant omicron cases in wave 5 with differences by vaccination status. This result is consistent with five other studies [9,17,20,21,28].

Furthermore, we found a much lower incidence of “loss of taste and smell”, which, in the past, was used as a test-triggering symptom [29,30], in the omicron cohort. These altogether increase the difficulty to distinguish COVID-19 with symptom-based testing algorithms. In fact, Fragaszy and colleagues calculated the specificity of such algorithms was as low as 47%-27% [31]. Although symptom-based information is of reduced value to diagnosis, we urge updating the symptom variation in public health messaging as failure to do so will hinder public awareness.

**Second**, the omicron variant may not be intrinsically less severe than preceding variants such that continued surveillance among vulnerable populations should remain in force. Unlike regions with a high incidence of natural infection prior to the omicron era, such as the United States [32], Hong Kong was almost free of infection in the community [13,14] coupled with low vaccine uptake rate among the old-old [2]. This allowed us to study the intrinsic severity of mortality among this disproportionately affected group. In contrast to Nyberg and colleagues who reported a larger reduction in the risk of death (from omicron to delta) among unvaccinated cases [15], we identified that the risk of death following SARS-CoV-2 infection among the unvaccinated elderly was similar between omicron and delta **(Table S1)**. We also found that omicron was more likely to cause fever than delta among unvaccinated symptomatic cases **(Table S3, Figure 2**). The reasons for the aforementioned contrasting findings need further investigation. In addition, when compared with other co-circulating respiratory illnesses, omicron, especially the BA.2 sublineage, is not intrinsically milder and can be more neuropathogenic [33].

**Third**, symptom-based predictions for death are possible and can help refine infection control policies to tackle COVID-19 in residential care homes for the elderly (RCHEs). Though a major surge in severe illnesses in RCHEs may be unlikely under a high vaccine or infection coverage [34], the delay in seeking medical care among the elderly [35], the multi-fold increase in mortality among institutionalized elderly [36], and the highest number of incidents attributable to RCHEs in the continual recurrent epidemics [6] altogether suggest active syndromic surveillance is needed in RCHEs. Further to the proposed frailty screening to be performed on every aged COVID-19 case to enable early risk stratification [37] and the inclusion of certain underlying health conditions as risk characteristics (such as hypertension [38], dyslipidemia and being bed-bound [39]), symptoms consistently predictive of severe clinical outcomes can be added to this individual-based risk assessment. Further to Ryan and colleagues who showed that severe COVID-19 could be predicted by symptoms in the wild-type era [40], we found certain symptoms (fever and shortness of breath) were robustly predictive of death during the wild-type-to-omicron transition. This finding is consistent with earlier reports in the wild-type era that shortness of breath was predictive of death [19,39].

**Fourth**, the stockpile of genus epidemicus should be optimized with the changing symptom profiles across vaccine doses given the long-term battle against COVID-19. In line with existing findings which reported changing symptom profiles across vaccine doses [9,28], we found that fever was less common as the number of vaccine doses increased, but some respiratory symptoms were still very common, including cough, runny nose, blocked nose and sore throat. Though the effect of vaccines on reducing severe outcomes following infection of COVID-19 is present across variants [41], it does not necessarily lower the occurrence of symptoms with high transmissibility. In line with an early report that virus shedding was initially highest in the upper respiratory tract [13], sore throat and runny nose were shown to be associated with higher viral load following infection with SARS-CoV-2 [17]. This suggests that the provision of medicine should be adjusted to suppress viral replication in human bodies with respect to the changing symptom profiles, particularly in regions with intense social-mixing like Hong Kong where the number of contacts reached 12.5 per day [42]. In Hong Kong, two antiviral agents (Molnupiravir and Paxlovid) were available to improve clinical outcomes, but their effect on symptom alleviation needs further research.

**Fifth**, future syndromic surveillance should be enriched by free-text reporting of symptoms by patients. By applying a multi-tier text-matching algorithm on free-text symptom data, we identified a wider disease spectrum, which exceeded the number of symptoms predefined by other studies [17,20,21].

Considering the four hard-coded symptoms in our dataset, we showed that a large proportion of identification was contributed by additional free-text data (3.8-47%), which is in line with Malden and colleagues who supplemented structured data of electronic medical records with natural language processing of unstructured data and found 29-64% additional cases for each selected symptoms [22].

### Study strengths and limitations

Clinical characteristics of SARS-CoV-2 were widely studied in the initial phase of the pandemic [43], but comparisons between recent subtypes were sparse and the intrinsic severity of subsequent variants were understudied. This is the first large-scale study which adds to the literature in this understudied area with free-text symptom reporting data from the patient perspective. In addition, unlike existing studies, which inferred the variant type to which a case belonged from the predominating strain circulating in the community at that time [17], we defined our cohort based on confirmed laboratory results, minimizing misclassification bias. However, there are **two** study limitations which bear mentioning. **First**, the duration and level of symptoms were not recorded such that we were unable to provide a better understanding of the difference between the same symptoms. **Second**, caution should be exercised when interpreting differences in symptom across variants, which may be biased by the potential underreporting of patient characteristics in the massive fifth omicron wave during which healthcare resources were stringent and by the changing method of data collection over time.

## Conclusion

Upon achieving herd immunity in the population to halt the exponential spread of SARS-CoV-2 [44], the next item on the agenda is to handle the disease burden caused by the changing symptom profile. Existing surveillance infrastructure should be updated with respect to changing symptomatology and mortality risk as SARS-CoV-2 and the population immunity level evolve. Such infrastructure can be bettered with reduced reliance on symptom-based case identification, increased weighing on symptoms robustly predictive of mortality in outcome prediction, strengthened infection control in RCHEs through universal individual-based risk assessment to enable early risk stratification, and adjusting the stockpile of medicine to target treatment arising from the changing clinical presentation against the background of changing immunity in the population. In addition, real-time algorithms to incorporate patients’ free-text reporting of symptoms should be developed to enrich syndromic surveillance.

## Data Availability

The availability of the data set is subject to approval from Hong Kong Centre for Health and Protection, Department of Health and relevant government departments.

## ACKNOWLEDGEMENT

We would like to acknowledge support from Health and Medical Research Fund (reference numbers: INF-CUHK-1, 17160302, 18170312, CID-CUHK-A, COVID1903008), General Research Fund (reference numbers: 14112818, 24104920), Wellcome Trust Fund (reference number: 200861/Z/16/Z) and Group Research Scheme of The Chinese University of Hong Kong. This work formed part of the doctoral thesis requirements for Wan In WEI.

## ETHICAL STATEMENT

This study was approved by the Survey and Behavioral Research Ethics Committee of The Chinese University of Hong Kong (reference: SBRE-19-595).

## AUTHORS’ CONTRIBUTIONS

Conceptualized: Kin On KWOK (KOK), Wan In WEI (WIW) Data curation: Edward B McNeil (EBM), WIW

Data analysis: EBM, WIW, KOK

First draft of the manuscript: WWI, KOK Data interpretation: KOK, EBM, WIW

Manuscript editing: EK YEOH (EKY), Samuel YS WONG (SYSW), EBM, WWI, KOK, Arthur TANG (AT), Julian TANG (JT)

Supervision: EKY, SYSW, KOK

Provision of critical comments: AT, SYSW, JT, EKY

