## Supplementary material for "Comparative analysis of symptom profile and risk of death associated with infection by SARS-CoV-2 and its variants in Hong Kong": Table S1; Table S2; Figure S1; Figure S2

**SUPPLEMENTARY MATERIALS**

**Table S1.** Methodological details on the iterative multi-tier text-matching algorithm

| Tier | Procedures |
|---|---|
| 1 | Text containing only words indicative of asymptomatic status were removed. We also standardized all Chinese Judou marks for easy identification of separate symptoms. All double quotes were replcaed with a single quote and doubles spaces with a single space. |
| 2 | Non-exhaustive removal of phrases fulfilling one of the following conditions:<br>● Text that induced subsequent false positive symptom mapping (identification)<br>● Text that mentioned existing health conditions or past medical history<br>● Symptoms that referred to persons other than the patient<br>● Symptoms that mentioned non-specific body parts<br>● Text gave unclear indication of any symptom<br>● Text unrelated to symptoms (for example, asking for sick leave certificate)<br>● Inconclusive phrases about body temperature<br>● Ambiguous body parts mentioned only<br>● Adjectives or generalizable conditions without any mention of body parts |
| 3 | Individual matching for phrases which were not suitable for global matching or contained patterns which were hard to generalize. For example, short-hand notations which caused false positive errors, and temperature descriptions ending with the cut-off digits. |
| 4 | Exact matching for fever or temperature with a clear indicator of minimum or maximum threshold |
| 5 | Exact matching for fever or temperature under the absence of specified phrases about a minimum or maximum threshold |
| 6 | Removal of negative descriptions, such as “no”, “not”, “don’t” and “afebrile” (and their Chinese equivalents) |
| 7 | Prioritized matching of selected symptom phrases to avoid subsequent false positive errors |
| 8 | Matching of symptom phrases under strict definitions |
| 9 | Matching of symptom phrases under less strict definitions |
| 10 | Matching of symptom phrases under strict definitions and under the following conditions:<br>● absence of certain phrases to avoid false positives; and/or<br>● matches a whole word (a standalone term). |
| 11 | Manual screening of the residual symptoms text to look up meaningful descriptions, which would then be used to update tiers 2-10 as appropriate. |

Remarks: Tiers 1-11 are iterative until no meaningful descriptions could be observed from the residual text in Tier 10. An anonymous sample dataset was uploaded to show the data structure of this study **[1]**.

**Table S2.** Prevalence of symptoms among COVID-19 cases stratified by vaccination status and variants. Symptoms with a combined prevalence of >0.5% are highlighted.

| | Unvaccinated | | | Vaccinated | |
|---|---|---|---|---|---|
| **Symptom** | **Wild type n= 1,175** | **Delta n= 95** | **Omicron (BA.2) n= 12,677** | **Delta n= 151** | **Omicron (BA.2) n= 54,129** |
| **General symptoms** | | | | | |
| Bodyache | 0.00 (0.00-0.33) | 0.00 (0.00-3.89) | 0.49 (0.38-0.63) | 0.00 (0.00-2.48) | 0.81 (0.74-0.89) |
| Chills | 2.13 (1.45-3.12) | 0.00 (0.00-3.89) | 1.82 (1.60-2.07) | 0.66 (0.12-3.66) | 1.93 (1.81-2.05) |
| Fatigue | 0.94 (0.52-1.67) | 2.11 (0.58-7.35) | 1.86 (1.64-2.11) | 1.99 (0.68-5.68) | 1.61 (1.50-1.71) |
| Fever | 37.2 (34.5-40.0) | 25.3 (17.6-34.8) | 36.2 (35.4-37.1) | 17.9 (12.6-24.8) | 19.2 (18.9-19.6) |
| Malaise | 2.47 (1.72-3.52) | 0.00 (0.00-3.89) | 0.09 (0.05-0.17) | 2.65 (1.03-6.61) | 0.12 (0.09-0.15) |
| Sweating | 0.09 (0.02-0.48) | 0.00 (0.00-3.89) | 0.05 (0.02-0.10) | 0.00 (0.00-2.48) | 0.04 (0.03-0.07) |
| **Nervous system** | | | | | |
| Confusion | 0.09 (0.02-0.48) | 0.00 (0.00-3.89) | 0.03 (0.01-0.08) | 0.00 (0.00-2.48) | 0.01 (0.00-0.02) |
| Convulsion | 0.00 (0.00-0.33) | 0.00 (0.00-3.89) | 0.01 (0.00-0.04) | 0.00 (0.00-2.48) | 0.00 (0.00-0.01) |
| Disturbance of taste and/or smell | 6.38 (5.12-7.93) | 1.05 (0.19-5.72) | 0.21 (0.15-0.31) | 6.62 (3.64-11.8) | 0.26 (0.22-0.31) |
| Dizziness | 0.77 (0.40-1.45) | 2.11 (0.58-7.35) | 1.05 (0.89-1.24) | 3.31 (1.42-7.52) | 0.84 (0.77-0.92) |
| Drowsiness | 0.00 (0.00-0.33) | 0.00 (0.00-3.89) | 0.02 (0.00-0.06) | 0.00 (0.00-2.48) | 0.03 (0.02-0.04) |
| Headache | 8.85 (7.36-10.6) | 13.7 (8.17-22.0) | 4.96 (4.60-5.35) | 14.6 (9.82-21.1) | 6.08 (5.89-6.29) |
| Insomnia | 0.00 (0.00-0.33) | 0.00 (0.00-3.89) | 0.03 (0.01-0.08) | 0.00 (0.00-2.48) | 0.03 (0.02-0.04) |
| Neuralgia | 0.00 (0.00-0.33) | 0.00 (0.00-3.89) | 0.00 (0.00-0.03) | 0.00 (0.00-2.48) | 0.01 (0.00-0.02) |
| Numbness | 0.00 (0.00-0.33) | 0.00 (0.00-3.89) | 0.02 (0.00-0.06) | 0.00 (0.00-2.48) | 0.01 (0.01-0.02) |
| Unsteadiness | 0.00 (0.00-0.33) | 0.00 (0.00-3.89) | 0.01 (0.00-0.04) | 0.00 (0.00-2.48) | 0.00 (0.00-0.01) |
| **Respiratory system** | | | | | |
| Blocked nose | 2.04 (1.38-3.02) | 0.00 (0.00-3.89) | 0.91 (0.76-1.09) | 3.97 (1.83-8.40) | 1.88 (1.76-1.99) |
| Breathing problem | 0.00 (0.00-0.33) | 0.00 (0.00-3.89) | 0.01 (0.00-0.04) | 0.00 (0.00-2.48) | 0.02 (0.01-0.04) |
| Breathing sound (stridor) | 0.00 (0.00-0.33) | 0.00 (0.00-3.89) | 0.01 (0.00-0.04) | 0.00 (0.00-2.48) | 0.00 (0.00-0.01) |
| Chest discomfort | 0.09 (0.02-0.48) | 1.05 (0.19-5.72) | 0.06 (0.03-0.11) | 0.66 (0.12-3.66) | 0.02 (0.01-0.04) |
| Chest pain | 0.43 (0.18-0.99) | 0.00 (0.00-3.89) | 2.03 (1.80-2.29) | 1.99 (0.68-5.68) | 2.49 (2.36-2.63) |
| Coryza (cold-like symptoms) | 0.00 (0.00-0.33) | 0.00 (0.00-3.89) | 0.48 (0.37-0.62) | 0.00 (0.00-2.48) | 0.91 (0.83-0.99) |
| Cough | 37.6 (34.9-40.4) | 38.9 (29.8-49.0) | 22.5 (21.8-23.3) | 47.0 (39.2-55.0) | 26.6 (26.2-27.0) |
| Influenza-like illness | 0.00 (0.00-0.33) | 0.00 (0.00-3.89) | 0.05 (0.02-0.10) | 0.00 (0.00-2.48) | 0.05 (0.04-0.07) |
| Nose bleed | 0.00 (0.00-0.33) | 0.00 (0.00-3.89) | 0.01 (0.00-0.04) | 0.00 (0.00-2.48) | 0.01 (0.00-0.02) |
| Nose dryness | 0.00 (0.00-0.33) | 0.00 (0.00-3.89) | 0.00 (0.00-0.03) | 0.00 (0.00-2.48) | 0.00 (0.00-0.01) |
| Pneumonia | 0.00 (0.00-0.33) | 0.00 (0.00-3.89) | 0.06 (0.03-0.11) | 0.00 (0.00-2.48) | 0.00 (0.00-0.01) |
| Respiratory tract infection | 0.00 (0.00-0.33) | 0.00 (0.00-3.89) | 0.02 (0.00-0.06) | 0.00 (0.00-2.48) | 0.02 (0.01-0.04) |
| Runny nose | 8.68 (7.20-10.4) | 20.0 (13.2-29.1) | 8.16 (7.70-8.65) | 33.1 (26.1-41.0) | 10.8 (10.5-11.0) |
| Shortness of breath | 5.19 (4.06-6.61) | 3.16 (1.08-8.88) | 3.73 (3.42-4.08) | 1.99 (0.68-5.68) | 3.84 (3.68-4.00) |
| Sinus pain | 0.00 (0.00-0.33) | 0.00 (0.00-3.89) | 0.00 (0.00-0.03) | 0.00 (0.00-2.48) | 0.00 (0.00-0.01) |
| Sneezing | 0.09 (0.02-0.48) | 0.00 (0.00-3.89) | 0.17 (0.11-0.25) | 0.00 (0.00-2.48) | 0.28 (0.24-0.33) |
| Sputum (expectoration) | 1.79 (1.17-2.72) | 3.16 (1.08-8.88) | 3.09 (2.80-3.41) | 1.99 (0.68-5.68) | 3.67 (3.52-3.83) |
| Voice disorders | 0.00 (0.00-0.33) | 0.00 (0.00-3.89) | 0.29 (0.21-0.40) | 0.00 (0.00-2.48) | 0.50 (0.44-0.56) |
| **Digestive system** | | | | | |
| Abdominal pain | 0.17 (0.05-0.62) | 0.00 (0.00-3.89) | 0.50 (0.40-0.64) | 0.00 (0.00-2.48) | 0.14 (0.11-0.17) |
| Constipation | 0.00 (0.00-0.33) | 0.00 (0.00-3.89) | 0.02 (0.01-0.07) | 0.00 (0.00-2.48) | 0.00 (0.00-0.01) |
| Diarrhea | 4.94 (3.84-6.33) | 4.21 (1.65-10.3) | 1.84 (1.62-2.09) | 0.66 (0.12-3.66) | 0.82 (0.75-0.90) |
| Dry mouth | 0.17 (0.05-0.62) | 0.00 (0.00-3.89) | 0.13 (0.08-0.21) | 0.66 (0.12-3.66) | 0.13 (0.10-0.16) |
| Dry throat | 0.17 (0.05-0.62) | 0.00 (0.00-3.89) | 0.38 (0.29-0.50) | 3.97 (1.83-8.40) | 0.79 (0.72-0.87) |
| Dysphagia | 0.00 (0.00-0.33) | 0.00 (0.00-3.89) | 0.00 (0.00-0.03) | 0.00 (0.00-2.48) | 0.01 (0.01-0.03) |

| | | | | | |
|---|---|---|---|---|---|
| Excessive saliva | 0.00 (0.00-0.33) | 0.00 (0.00-3.89) | 0.00 (0.00-0.03) | 0.00 (0.00-2.48) | 0.00 (0.00-0.01) |
| Itchy throat | 0.09 (0.02-0.48) | 2.11 (0.58-7.35) | 0.43 (0.33-0.56) | 1.99 (0.68-5.68) | 0.88 (0.80-0.96) |
| Lip swelling | 0.00 (0.00-0.33) | 0.00 (0.00-3.89) | 0.00 (0.00-0.03) | 0.00 (0.00-2.48) | 0.00 (0.00-0.01) |
| Loss of appetite | 0.17 (0.05-0.62) | 0.00 (0.00-3.89) | 0.35 (0.26-0.47) | 0.00 (0.00-2.48) | 0.06 (0.05-0.09) |
| Mouth discomfort | 0.00 (0.00-0.33) | 0.00 (0.00-3.89) | 0.00 (0.00-0.03) | 0.00 (0.00-2.48) | 0.00 (0.00-0.01) |
| Mouth pain | 0.00 (0.00-0.33) | 0.00 (0.00-3.89) | 0.04 (0.02-0.09) | 0.00 (0.00-2.48) | 0.03 (0.02-0.05) |
| Nausea or vomiting | 0.68 (0.35-1.34) | 1.05 (0.19-5.72) | 2.54 (2.28-2.83) | 1.32 (0.36-4.70) | 0.51 (0.46-0.58) |
| Sore throat | 15.0 (13.1-17.1) | 18.9 (12.3-28.0) | 16.0 (15.3-16.6) | 29.1 (22.5-36.8) | 25.0 (24.6-25.4) |
| Throat discomfort | 1.53 (0.97-2.41) | 1.05 (0.19-5.72) | 0.28 (0.21-0.39) | 0.66 (0.12-3.66) | 0.59 (0.53-0.66) |
| Throat irritation | 0.00 (0.00-0.33) | 0.00 (0.00-3.89) | 0.00 (0.00-0.03) | 0.00 (0.00-2.48) | 0.01 (0.00-0.02) |
| Tongue pain | 0.00 (0.00-0.33) | 0.00 (0.00-3.89) | 0.00 (0.00-0.03) | 0.00 (0.00-2.48) | 0.01 (0.00-0.02) |
| Tonsillitis | 0.00 (0.00-0.33) | 0.00 (0.00-3.89) | 0.02 (0.00-0.06) | 0.00 (0.00-2.48) | 0.01 (0.01-0.03) |
| Toothache | 0.00 (0.00-0.33) | 0.00 (0.00-3.89) | 0.00 (0.00-0.03) | 0.00 (0.00-2.48) | 0.01 (0.01-0.02) |
| **Circulatory system** | | | | | |
| Collapse | 0.09 (0.02-0.48) | 0.00 (0.00-3.89) | 0.00 (0.00-0.03) | 0.00 (0.00-2.48) | 0.00 (0.00-0.01) |
| Heart discomfort | 0.00 (0.00-0.33) | 0.00 (0.00-3.89) | 0.00 (0.00-0.03) | 0.00 (0.00-2.48) | 0.00 (0.00-0.01) |
| Irregular blood pressure | 0.00 (0.00-0.33) | 0.00 (0.00-3.89) | 0.02 (0.01-0.07) | 0.00 (0.00-2.48) | 0.02 (0.01-0.04) |
| Lymphadenopathy | 0.00 (0.00-0.33) | 0.00 (0.00-3.89) | 0.00 (0.00-0.03) | 0.00 (0.00-2.48) | 0.00 (0.00-0.01) |
| Palpitations | 0.00 (0.00-0.33) | 0.00 (0.00-3.89) | 0.02 (0.01-0.07) | 0.66 (0.12-3.66) | 0.02 (0.01-0.04) |
| Tachycardia | 0.00 (0.00-0.33) | 0.00 (0.00-3.89) | 0.05 (0.02-0.10) | 0.00 (0.00-2.48) | 0.03 (0.02-0.05) |

| | | | | | |
|---|---|---|---|---|---|
| **Musculoskeletal system** | | | | | |
| Back pain | 0.26 (0.09-0.75) | 0.00 (0.00-3.89) | 0.21 (0.15-0.31) | 0.66 (0.12-3.66) | 0.20 (0.16-0.24) |
| Face and/or neck pain | 0.00 (0.00-0.33) | 0.00 (0.00-3.89) | 0.02 (0.01-0.07) | 0.00 (0.00-2.48) | 0.01 (0.01-0.03) |
| Face and/or neck stiffness | 0.00 (0.00-0.33) | 0.00 (0.00-3.89) | 0.01 (0.00-0.04) | 0.00 (0.00-2.48) | 0.00 (0.00-0.01) |
| Face and/or neck swelling | 0.00 (0.00-0.33) | 0.00 (0.00-3.89) | 0.00 (0.00-0.03) | 0.00 (0.00-2.48) | 0.00 (0.00-0.01) |
| Lowerextremity coldness | 0.00 (0.00-0.33) | 0.00 (0.00-3.89) | 0.01 (0.00-0.04) | 0.00 (0.00-2.48) | 0.02 (0.01-0.03) |
| Lowerextremity cramp | 0.00 (0.00-0.33) | 0.00 (0.00-3.89) | 0.01 (0.00-0.04) | 0.00 (0.00-2.48) | 0.00 (0.00-0.01) |
| Lowerextremity pain | 0.00 (0.00-0.33) | 0.00 (0.00-3.89) | 0.13 (0.08-0.21) | 0.00 (0.00-2.48) | 0.09 (0.07-0.12) |
| Lowerextremity swelling | 0.09 (0.02-0.48) | 0.00 (0.00-3.89) | 0.00 (0.00-0.03) | 0.00 (0.00-2.48) | 0.00 (0.00-0.01) |
| Ostalgia | 0.17 (0.05-0.62) | 0.00 (0.00-3.89) | 0.45 (0.35-0.58) | 0.00 (0.00-2.48) | 0.70 (0.64-0.78) |
| Unspecified joint pain | 0.09 (0.02-0.48) | 0.00 (0.00-3.89) | 0.21 (0.15-0.31) | 0.00 (0.00-2.48) | 0.14 (0.11-0.17) |
| Unspecified joint stiffness | 0.00 (0.00-0.33) | 0.00 (0.00-3.89) | 0.00 (0.00-0.03) | 0.00 (0.00-2.48) | 0.00 (0.00-0.01) |
| Unspecified limb pain | 0.00 (0.00-0.33) | 0.00 (0.00-3.89) | 0.00 (0.00-0.03) | 0.00 (0.00-2.48) | 0.00 (0.00-0.01) |
| Unspecified muscle pain | 2.81 (2.01-3.92) | 1.05 (0.19-5.72) | 1.59 (1.38-1.82) | 3.31 (1.42-7.52) | 1.92 (1.80-2.03) |
| Unspecified muscle stiffness | 0.00 (0.00-0.33) | 0.00 (0.00-3.89) | 0.00 (0.00-0.03) | 0.00 (0.00-2.48) | 0.00 (0.00-0.01) |
| Upperextremity coldness | 0.00 (0.00-0.33) | 0.00 (0.00-3.89) | 0.01 (0.00-0.04) | 0.00 (0.00-2.48) | 0.02 (0.01-0.03) |
| Upperextremity cramp | 0.00 (0.00-0.33) | 0.00 (0.00-3.89) | 0.01 (0.00-0.04) | 0.00 (0.00-2.48) | 0.00 (0.00-0.01) |
| Upperextremity pain | 0.00 (0.00-0.33) | 0.00 (0.00-3.89) | 0.06 (0.03-0.12) | 0.00 (0.00-2.48) | 0.09 (0.07-0.12) |
| Weak joint(s) | 0.00 (0.00-0.33) | 0.00 (0.00-3.89) | 0.00 (0.00-0.03) | 0.00 (0.00-2.48) | 0.01 (0.00-0.02) |
| Weak limb(s) | 0.00 (0.00-0.33) | 0.00 (0.00-3.89) | 0.14 (0.09-0.22) | 0.00 (0.00-2.48) | 0.17 (0.14-0.21) |
| Weak muscle(s) | 0.00 (0.00-0.33) | 0.00 (0.00-3.89) | 0.01 (0.00-0.04) | 0.00 (0.00-2.48) | 0.02 (0.01-0.03) |
| **Urinary system** | | | | | |
| Ambiguous urine problems | 0.00 (0.00-0.33) | 0.00 (0.00-3.89) | 0.00 (0.00-0.03) | 0.00 (0.00-2.48) | 0.00 (0.00-0.01) |
| Frequent urination | 0.00 (0.00-0.33) | 0.00 (0.00-3.89) | 0.01 (0.00-0.04) | 0.00 (0.00-2.48) | 0.00 (0.00-0.01) |
| Incontinence of urine | 0.00 (0.00-0.33) | 0.00 (0.00-3.89) | 0.00 (0.00-0.03) | 0.00 (0.00-2.48) | 0.00 (0.00-0.01) |
| Painful urination | 0.00 (0.00-0.33) | 0.00 (0.00-3.89) | 0.01 (0.00-0.04) | 0.00 (0.00-2.48) | 0.00 (0.00-0.01) |
| Retention of urine | 0.00 (0.00-0.33) | 0.00 (0.00-3.89) | 0.00 (0.00-0.03) | 0.00 (0.00-2.48) | 0.00 (0.00-0.01) |
| Scrotum pain | 0.00 (0.00-0.33) | 0.00 (0.00-3.89) | 0.01 (0.00-0.04) | 0.00 (0.00-2.48) | 0.00 (0.00-0.01) |
| **Other** | | | | | |
| Ambiguous ear problems | 0.00 (0.00-0.33) | 0.00 (0.00-3.89) | 0.01 (0.00-0.04) | 0.00 (0.00-2.48) | 0.00 (0.00-0.01) |
| Anxiety | 0.00 (0.00-0.33) | 0.00 (0.00-3.89) | 0.00 (0.00-0.03) | 0.00 (0.00-2.48) | 0.00 (0.00-0.01) |
| Blisters | 0.00 (0.00-0.33) | 0.00 (0.00-3.89) | 0.00 (0.00-0.03) | 0.00 (0.00-2.48) | 0.00 (0.00-0.01) |
| Blocked ear(s) | 0.00 (0.00-0.33) | 0.00 (0.00-3.89) | 0.01 (0.00-0.04) | 0.00 (0.00-2.48) | 0.00 (0.00-0.01) |
| Blurred vision | 0.00 (0.00-0.33) | 0.00 (0.00-3.89) | 0.01 (0.00-0.04) | 0.00 (0.00-2.48) | 0.01 (0.00-0.02) |

| | | | | | | |
|---|---|---|---|---|---|---|
| Eye discharge | 0.00 (0.00-0.33) | 0.00 (0.00-3.89) | 0.00 (0.00-0.03) | | 0.00 (0.00-2.48) | 0.02 (0.01-0.03) |
| Eye dryness | 0.00 (0.00-0.33) | 0.00 (0.00-3.89) | 0.00 (0.00-0.03) | | 0.00 (0.00-2.48) | 0.00 (0.00-0.01) |
| Eye itchiness | 0.00 (0.00-0.33) | 0.00 (0.00-3.89) | 0.00 (0.00-0.03) | | 0.00 (0.00-2.48) | 0.00 (0.00-0.01) |
| Eye lid issues | 0.00 (0.00-0.33) | 0.00 (0.00-3.89) | 0.00 (0.00-0.03) | | 0.00 (0.00-2.48) | 0.00 (0.00-0.01) |
| Eye pain | 0.00 (0.00-0.33) | 0.00 (0.00-3.89) | 0.02 (0.01-0.07) | | 0.00 (0.00-2.48) | 0.02 (0.01-0.04) |
| Feeling low | 0.00 (0.00-0.33) | 0.00 (0.00-3.89) | 0.00 (0.00-0.03) | | 0.00 (0.00-2.48) | 0.00 (0.00-0.01) |
| Hearing dysfunction | 0.00 (0.00-0.33) | 0.00 (0.00-3.89) | 0.02 (0.00-0.06) | | 0.00 (0.00-2.48) | 0.03 (0.02-0.05) |
| Itchy ear(s) | 0.00 (0.00-0.33) | 0.00 (0.00-3.89) | 0.00 (0.00-0.03) | | 0.00 (0.00-2.48) | 0.00 (0.00-0.01) |
| Rash | 0.00 (0.00-0.33) | 0.00 (0.00-3.89) | 0.06 (0.03-0.11) | | 0.00 (0.00-2.48) | 0.03 (0.02-0.05) |
| Red eye(s) | 0.00 (0.00-0.33) | 0.00 (0.00-3.89) | 0.02 (0.00-0.06) | | 0.00 (0.00-2.48) | 0.01 (0.01-0.02) |
| Skin irritation | 0.00 (0.00-0.33) | 0.00 (0.00-3.89) | 0.01 (0.00-0.04) | | 0.00 (0.00-2.48) | 0.01 (0.00-0.02) |
| Sore ear(s) | 0.00 (0.00-0.33) | 0.00 (0.00-3.89) | 0.05 (0.02-0.10) | | 0.00 (0.00-2.48) | 0.06 (0.05-0.09) |

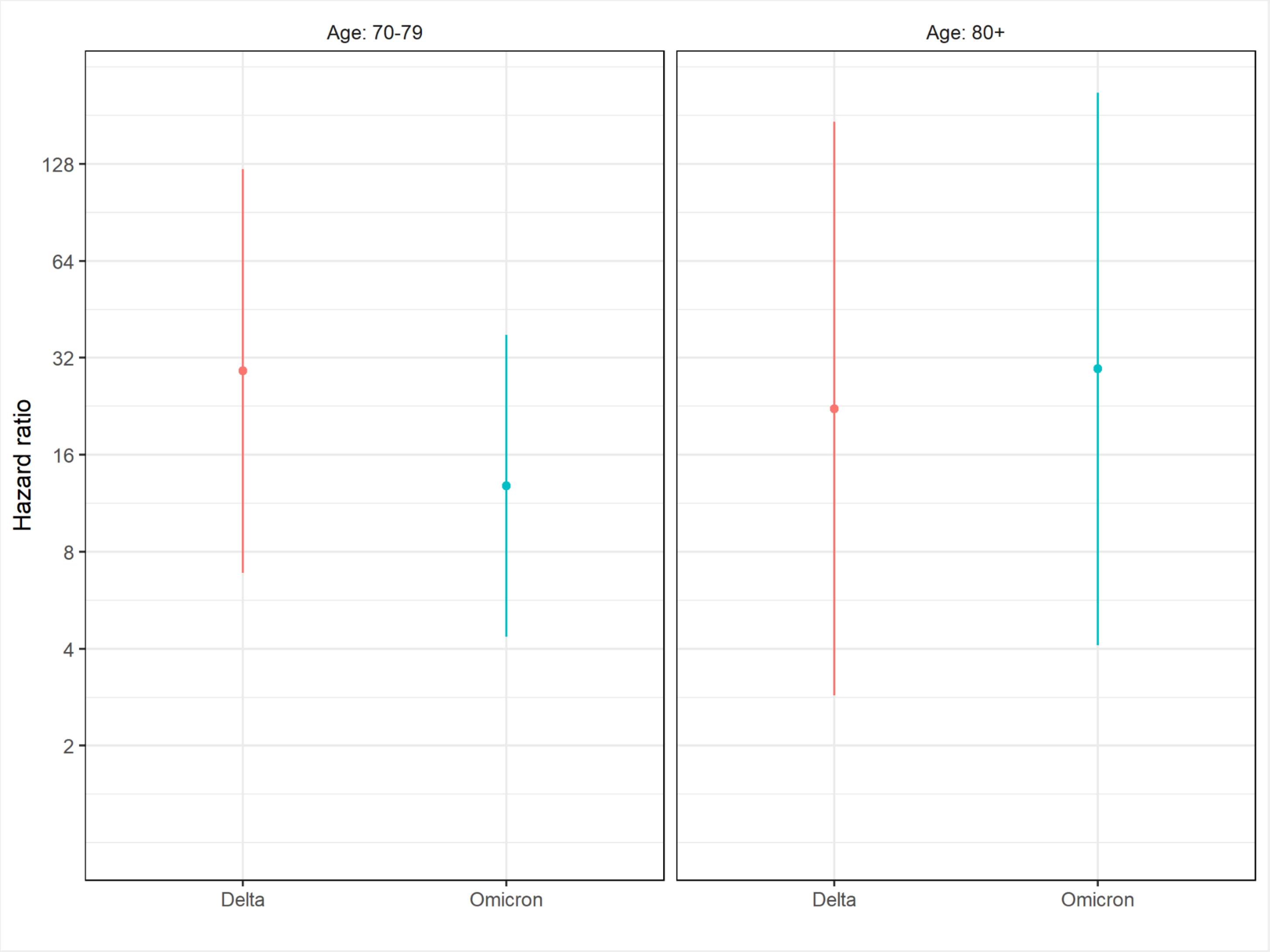


**Figure S1**. Adjusted hazard ratios of death within 28 days of onset for delta and omicron variants versus the wild-type among unvaccinated elderly cases aged 70 years or above.

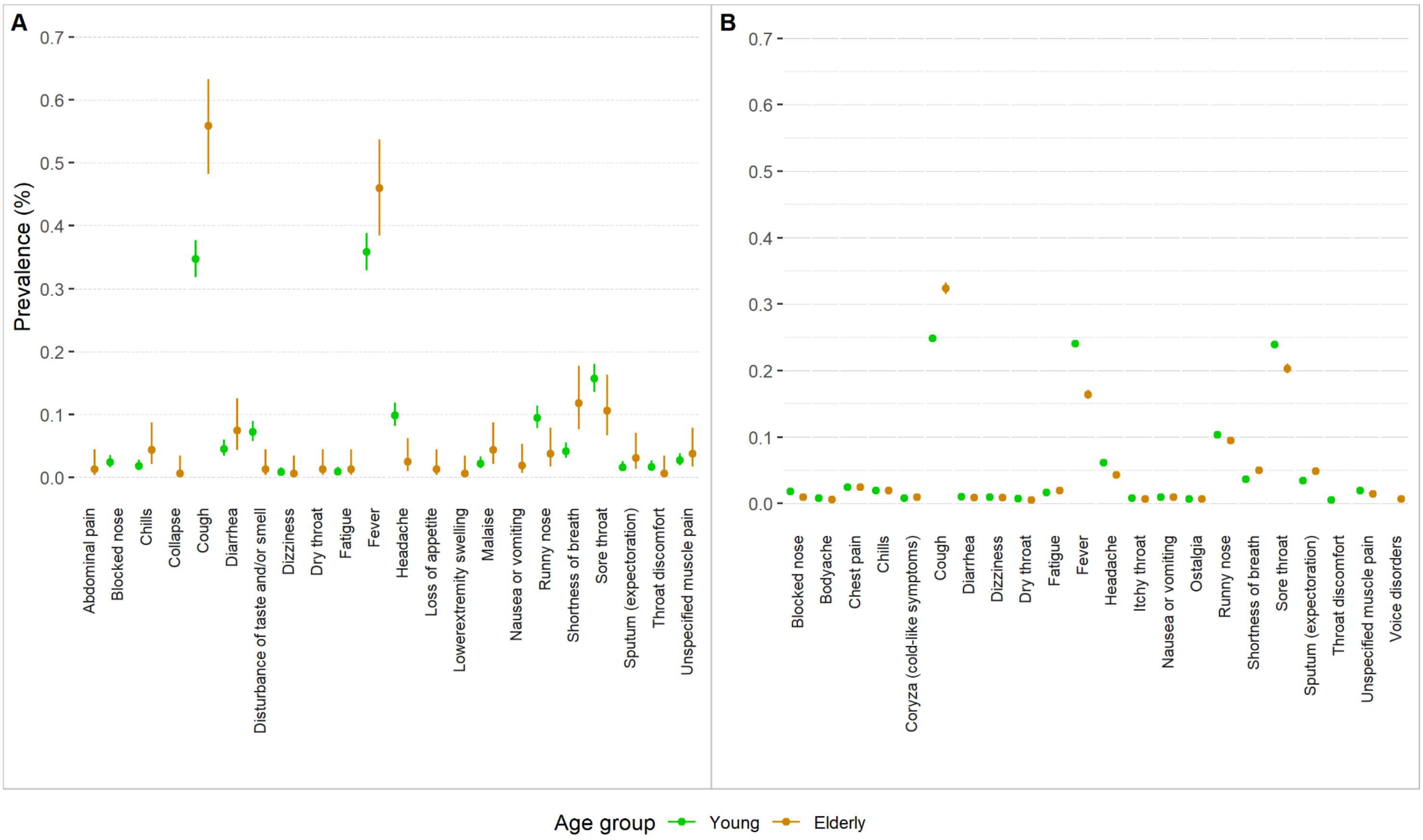


**Figure S2**. Prevalence of common symptoms among unvaccinated cases stratified by age group for (A) the wild type and (B) omicron BA.2 sub-variant. Vertical segments are 95% confidence intervals.
